# Dose-Escalation Study of Amygdalar Transcranial Focused Ultrasound in Healthy Volunteers

**DOI:** 10.1101/2025.10.21.25338018

**Authors:** Norman M. Spivak, Andrew A.E.D. Bishay, Jonathan Haroon, Amber R. Hopkins, Jody Tanabe, Sabrina Halavi, Bianca Hoang-Dang, Andrew J. Swenson, Samantha F. Schafer, Mark E. Schafer, Alexander Bystritsky, Susan Y. Bookheimer, Martin M. Monti

**Author notes:** Senior Author (Martin M. Monti).

## Abstract

**Background:** Transcranial focused ultrasound (tFUS) is a promising noninvasive technique for modulating deep brain structures, but the optimal and safe intensity range for neuromodulation remains unclear. Current FDA intensity limits, designed for diagnostic use rather than therapy, may limit the potential effectiveness of tFUS.

**Objective:** This study aimed to evaluate the safety and preliminary efficacy of escalating tFUS intensities targeting the right amygdala in healthy volunteers, including intensity levels exceeding current FDA diagnostic ultrasound limits.

**Methods:** Ten healthy adults (mean age = 19.5 ± 1.4 years; 70% female) participated in a within-subject, repeated-measures design. Each received four randomized tFUS stimulation conditions (ISPTA.3 = 0.72–10.08 W/cm²) using the BX Pulsar 1002 system. Structural MRI was performed before each stimulation to monitor safety. Affective modulation was assessed pre- and post-stimulation via the International Affective Picture Set (IAPS) task, measuring changes in valence and arousal ratings. Acoustic modeling was conducted using BabelBrain to estimate intracranial energy deposition.

**Results:** No adverse events or MRI-detectable structural changes (e.g., edema, hemorrhage, or microstructural abnormalities) occurred under any stimulation condition. Behavioral analyses revealed no significant overall effects of intensity on affective measures. Comparisons between excitatory and inhibitory paradigms showed no significant differences.

**Conclusions:** tFUS of the right amygdala at intensities up to 10.08 W/cm² ISPTA.3—over 14 times the FDA diagnostic limit—was well tolerated and produced no structural or subjective adverse effects. These findings support the safety of higher-intensity tFUS and justify further investigation in larger and clinical populations to explore potential dose–response effects on emotion regulation.

## Introduction

Determining the optimal intensity of transcranial focused ultrasound (tFUS) needed to effectuate change in neural activity is a critical step in moving towards using tFUS as a safe and effective neuromodulatory tool. While prior studies have established the feasibility of modulating deep brain structures such as the amygdala, thalamus, and entorhinal cortex, these all operated within the Food and Drug Administration’s (FDA) output limit for diagnostic ultrasound [1–4].

In 1985, the United States FDA established exposure limits for diagnostic ultrasound (peripheral vascular, 720 mW/cm^2^; cardiac, 430 mW/cm^2^; fetal and other, 94 mW/cm^2^; and ophthalmic, 17 mW/cm^2^ ; all are derated intensity spatial peak temporal average (I_SPTA.3_)) on the basis of the highest intensities being used when the FDA gained regulatory control. In cooperation with the ultrasonic community, these values were revised in 1992, establishing a new limit of I_SPTA.3_ = 720 mW/cm^2^ for all applications except those involving ophthalmic applications. The vast majority of tFUS work uses the FDA "Track 3" requirements, which require the following: I_SPTA.3_ must not exceed 720 mW/cm^2^, and either the Mechanical Index (MI) must be below 1.9 or the intensity spatial peak pulse average (I_SPPA.3_) must be below 190 W/cm^2^. However, these limits apply to diagnostic ultrasound, not therapeutic ultrasound. Nevertheless, many researchers abide by these limits for ease of gaining approval from oversight bodies (such as Institutional Review Boards (IRB)) to conduct their research. Furthermore, many publications over the last two decades have underscored both the safety record of tFUS [5–8] and its ability to exert non-invasively neuromodulatory effects[9, 10]. It is important to note that these guidelines do not imply that these are the safety limits, above which stimulation would be unsafe and that stimulation below these intensities is automatically safe.

While the FDA limit has allowed for ease in the obtaining of regulatory approval from an IRB to proceed with these studies, neuromodulation requires different considerations, as ultrasound delivered at intensities compliant with the limit may evoke non-uniform responses attributable to the stochastic nature of neuronal firing. Accordingly, variability at FDA-compliant outputs should not be taken to imply a lack of efficacy; rather, it suggests that achieving reliable neuromodulatory engagement may require operation within a parameter space not fully encompassed by diagnostic ultrasound limits. In fact, multiple studies have shown the safety of intensities exceeding the FDA limits[11–14]. Our group conducted an open-label study of ultrasound administration to the epileptogenic focus in pre-surgical temporal lobe epilepsy patients who subsequently underwent resection of the epileptogenic zone to treat their refractory epilepsy. In that work, we did not see histological changes in brain tissue when sonicating up to I_SPTA.3_ of 5.76 W/cm^2^ (eight times above the FDA limit)[13]. However, since in-vivo sonication requires transmission through the skin and skull, which inherently causes attenuation and some loss of energy[15], we aimed to first understand the minimum energy required to cause damage in tissue directly. An *ex vivo* brain tissue sample study was designed to determine safety thresholds for ultrasonic brain stimulation. In that work, pieces of brain tissue from recently deceased cadavers and surgical resections were directly sonicated in a petri dish at varying intensity levels to determine the threshold at which damage starts to appear on histology[11]. No signs of damage were observed in any samples that underwent sonication at or below 14 W/cm² I_SPTA.0_, while inconsistent damage above this intensity included changes such as extravasation of blood vessels and spongiosis. Again, there was no skull to provide for any attenuation of ultrasonic energy. While the aforementioned study involved direct *ex-vivo* sonication of brain tissue, studies in living humans involve transcranial administration through skin, skull, and hair. It was therefore important to understand if the increased energy deposition associated with higher intensity stimulation (and subsequent energy absorption by the skull) would result in appreciable heating of the skin and skull. A small study in healthy volunteers involved sonication of the right scapula, administered from the dorsal surface[16]. With thermoprobes placed between the ultrasound transducer and the skin, the temperature was measured during stimulation with five different intensity levels up to 14 W/cm² I_SPTA.3_. Even at the maximum stimulation intensity, only 1.5° C of heating were seen, which is a reasonable rise in temperature that is not expected to produce adverse events and which is below the FDA cutoff of 2° C. These findings thus suggest the presence of a safe window of increased energy deposition well above the current FDA energy limit, implying a region of ultrasound parametrization capable of increased neuromodulation and, putatively, increased therapeutic effectiveness, without a concomitant increase in risk profile.

The possibility of employing intensities above the FDA limit is particularly important, since the skull substantially attenuates ultrasound[17], which may greatly limit the effect sizes induced by ultrasonic neuromodulation. As such, at intensities compliant with the FDA limit, there is limited energy delivered to the tissue itself, which limits the neuromodulatory effect. This raises a central question: does an intensity range above the FDA limit permit stronger neuromodulatory effects without compromising safety, and is there a dose–response such that increasing intensity yields more reliable or larger changes in brain activity?

The present study aims to fill this gap through a carefully designed dose-escalation study conducted in healthy volunteers. Specifically, using four carefully chosen stimulation paradigms, we tested whether tFUS of the right amygdala at intensities above the FDA limit had any differential effect on a behavioral task previously validated with amygdalar tFUS[1]. It is known that the amygdala plays a role in various emotional and behavioral responses, and that amygdalar circuits are crucial for anxiety and fear processing and play an essential role in regulating emotions[18, 19]. Given the significant role of the amygdala in emotional processing and its links to psychiatric disorders, investigating clinical interventions aimed at modulating amygdalar activity is key to making forward therapeutic progress. We also acquired clinical MR sequences to monitor for changes indicative of injury caused by tFUS[20]. We hypothesized that higher stimulation intensities would lead to more significant emotional shifts as assessed by changes in the behavioral tasks, while ensuring that the increased intensities would not result in tissue injury as assessed by structural MRI.

## Materials and Methods

### Participants

Ten healthy volunteers (age (SD): 19.5 (1.4) years; 70% female) were recruited and screened for eligibility. Inclusion required normal or corrected-to-normal vision and hearing. Participants with any DSM-5 diagnosis, current use of psychiatric medications, MRI contraindications (such as implants), or pregnancy were excluded. The study was reviewed by the UCLA Medical IRB # 3 (IRB-23-0612). All subjects signed a written informed consent form prior to participating in any study activities. Subjects were compensated $50 per study visit with an additional $50 completion bonus, for a total compensation of $300 for completing all five study visits.

### Study Design

This was a within-subjects, repeated-measures design in which each subject received four different tFUS stimulation parametrizations—randomized and counterbalanced across visits—to the right amygdala. Each stimulation visit also required subjects to complete a behavioral task twice, once before and once after stimulation, as well as undergo a structural MR acquisition, which was always performed before tFUS administration. There was a fifth follow-up visit with imaging only. Subjects were prospectively queried for changes in adverse events suggested by Legon and colleagues[21]. All study visits were spaced one week apart. The tFUS parameters used are shown in **Table 1**.

**Table 1:** tFUS Stimulation Parameters.

| Parameter Set | ISPTA.3 (W/cm<sup>2</sup>) | ISPPA.3 (W/cm<sup>2</sup>) | Pr.3 (MPa) | Mechanical Index | Duty Cycle (%) | Pulse Repetition Frequency (Hz) | Pulse Duration (ms) |
| --- | --- | --- | --- | --- | --- | --- | --- |
| A | 0.72 | 1.03 | 0.177 | 0.220 | 70 | 1000 | 0.7 |
| B | 5.76 | 8.23 | 0.477 | 0.592 | 70 | 1000 | 0.7 |
| C | 10.08 | 14.4 | 0.635 | 0.788 | 70 | 1000 | 0.7 |
| D | 10.08 | 201.6 | 2.26 | 2.803 | 4.97 | 71 | 0.7 |

Parameters were chosen as follows: In all paradigms, the pulse length was held constant at 0.7 ms. Parameter Set A uses the FDA output limit for the derated spatial-peak temporal-average intensity, with a 70% duty cycle, which is posited to be excitatory by Yoon et al [12] and likewise suggested in a review paper by Dell’Italia and colleagues[22]. Parameter Set A was then modified to have a derated spatial-peak temporal-average intensity that matches the maximum intensity used in a safety study by Stern and colleagues to yield Parameter Set B[13]. Parameter Set C was a modification of Parameter Set B with a derated spatial-peak pulse-average intensity of 14.4 W/cm^2^ that matches that of Cain and colleagues[23]. Lastly, Parameter Set D was made by modifying Parameter Set C to maintain the same derated spatial-peak temporal-average intensity but with a lower duty cycle at 5%, which Dell’Italia suggests as being inhibitory. In this case, Parameter Set 4 was at 4.97% because of the need to maintain a 0.7 ms pulse length. (N.B., We do not believe that a difference of 0.03% is biologically meaningful.)

### Focused Ultrasound Device

The BX Pulsar 1002 Focused Ultrasound Device (BrainSonix Corp, Sherman Oaks, CA) was used to deliver the stimulation in this study[24, 25]. The BX Pulsar 1002 operates at a fundamental frequency of 650 kHz and uses transducers with a 61.5 mm aperture diameter, with varying 35 radii of curvature to allow for different focal depths of stimulation. In this study, we used either the 55 or 65mm focal depth transducer, depending on individual neuroanatomy, which varied per subject. The transducer was coupled to the subject’s temporal window using BrainSonix transmit acoustic standoff pads and sufficient ultrasound gel (Aquasonic 100, Parker Laboratories) to wet the hair and ensure no air bubbles would be present that could potentially attenuate ultrasound.

### Imaging and Neuronavigation

All MRI scans were acquired on a Siemens 3T Prisma scanner using a 32 channel coil at UCLA’s Staglin Center for Cognitive Neuroscience. Each session included a high-resolution T1-weighted MPRAGE scan for anatomical localization (TR/TE = 2000/2.52 ms, TI = 1100 ms, flip angle = 12°, voxel size = 1 mm³ isotropic, GRAPPA = 2, 192 slices), a T2-weighted turbo spin echo (TR/TE = 5000/104 ms, flip angle = 120°, voxel size = 0.5 × 0.5 × 4.0 mm³, GRAPPA = 2, 25 slices), and a 3D susceptibility-weighted imaging (SWI) scan sensitive to venous vasculature and microhemorrhages (TR/TE = 27/20 ms, flip angle = 15°, voxel size = 0.9 × 0.9 × 1.5 mm³, GRAPPA = 2, 80 slices). Diffusion imaging was acquired with a RESOLVE sequence for distortion-reduced diffusion imaging (TR = 4050 ms, TE1/TE2 = 63/111 ms, flip angle = 180°, voxel size = 0.5 × 0.5 × 4.0 mm³, 4 directions, 27 slices). All four sequences were repeated at each session to monitor for structural changes potentially resulting from the prior week’s ultrasound stimulation. An expert neuroradiologist reviewed the first and final session scans, with the intermediate scans scheduled for review only if changes were observed.

Additionally, the T1-weighted image from the first session was then subsequently used at every session for neuronavigation with BrainSight (Rogue Research, Montreal, QC), which allowed for positioning of the transducer aimed at the right amygdala without the need for real-time MR guidance.

### Acoustic Modeling

Acoustic modeling was performed using BabelBrain[26]. BabelBrain allows for the simulation of acoustic effects achieved with an ultrasound transducer trajectory (location and orientation of the transducer toward a target region) for an individual brain, considering the type of transducer used, the distance from the transducer surface to the scalp, the sonication parameters, as well as distortion effects caused by the skull (estimated from the individual T1-weighted data). The fixed-focal-length Brainsonix transducer used in this work (65-mm) is supported in BabelBrain. We selected one random participant per parameter set for modeling to demonstrate how the acoustic effects achieved differed between the parameter sets, as well as how the simulated focus of the ultrasound beam compared across participants, considering differences in transducer position and individual head characteristics. Skull and tissue characteristics estimated by BabelBrain and considered when simulating the acoustic effects achieved for each selected participant are presented in Fig. 1.

**Figure 1:**
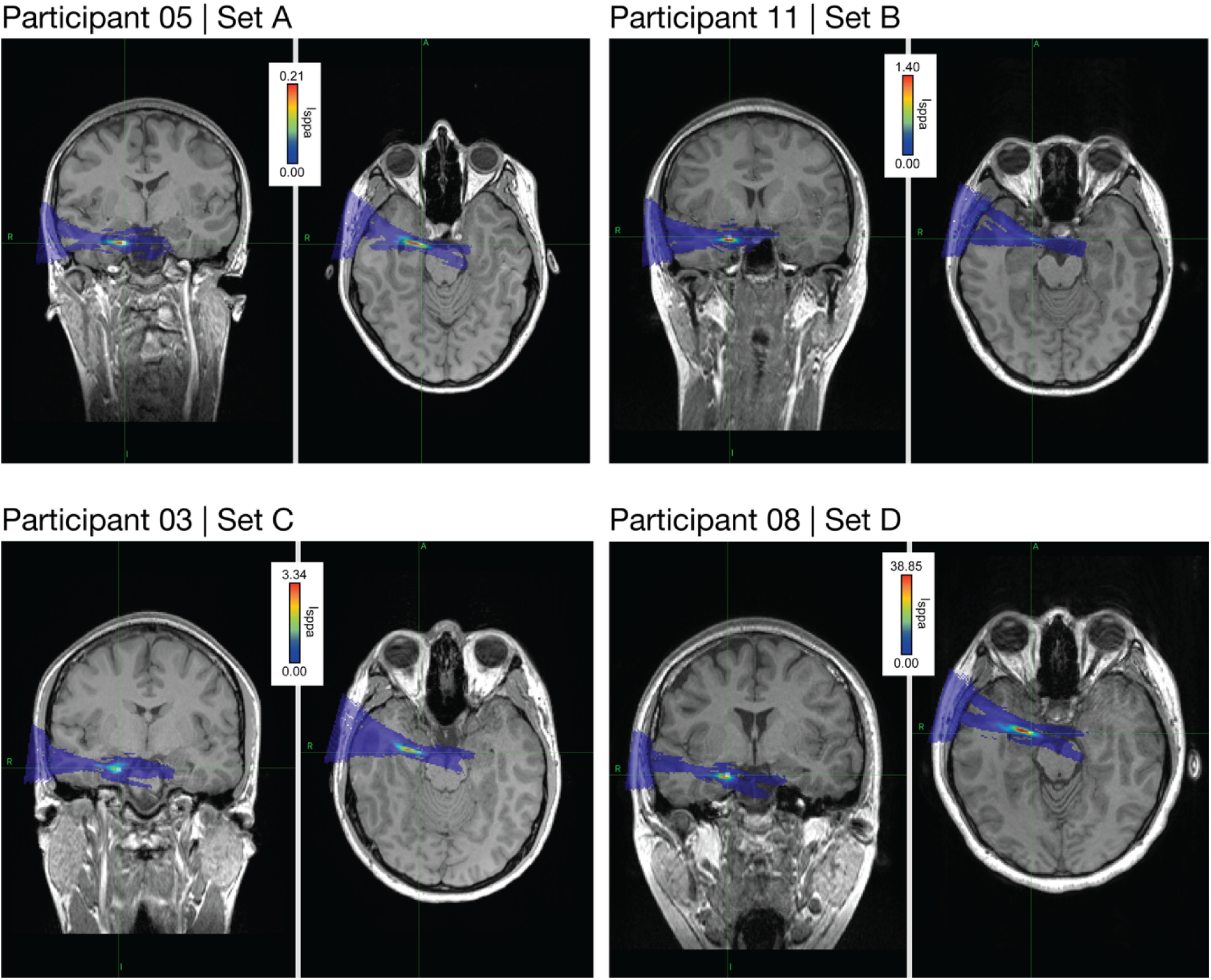
Simulated acoustic effects of sonication with each parameter set. One participant was selected for modeling each parameter set. Green crosshairs intersect at the single voxel coordinate defined in the neuronavigational system that tFUS was aimed at for each participant. A target region (3×3×3 mm white cube) was generated around this single coordinate to better compare the simulated acoustic effects against the target region. The focus of the simulated ultrasound beam is colored by the estimated acoustic pressure achieved, ranging least (dark blue) to greatest (dark red) pressure achieved for the individual brain.

### Behavioral Task

Before and after each stimulation, subjects completed a computer task using the International Affective Picture Set (IAPS)[1, 27]. Here, the subjects rated the valence and arousal of emotionally salient images, and changes in arousal and valence were used as a proxy for modulation of amygdalar-driven affective activity. There were eight different forms of the task, for which the images were randomized and counterbalanced among the sessions, and the forms themselves were designed to have balanced normative means for arousal and valence. Furthermore, each form of the task had balanced normative means for valence of three subcategories of images: positive, neutral, and negative[28, 29].

To assess stimulation effects on affective responses, we calculated within-subject pre- /post-task differences in positive and negative valence ratings for each stimulation parameter set. Additionally, a directional valence index (DVI) was computed for each condition as the change in positive valence minus the absolute change in negative valence (ΔPositive − |ΔNegative|). Likewise, a directional arousal Index (DAI) was computed for each condition as the change in positive arousal minus the absolute change in negative arousal (ΔPositive − |ΔNegative|). Normality of the arousal and valence difference distributions, DAI, and DVI distribution was assessed via the Shapiro–Wilk test. To assess differences in the arousal and valence differences, DAI, and DVI across stimulation paradigms, we used a repeated measures ANOVA with post-hoc pairwise comparisons with Holm correction for multiple testing as needed. For deviations from normality, a Wilcoxon signed-rank test or a Friedman test was used as appropriate. As Parameter Set D is inhibitory, arousal and valence changes, DAI, and DVI were analyzed against Parameter Set C, as they have equivalent I_SPTA.3_ levels to see if there is a directional effect using a paired t-test. Arousal and valence differences, DAI, and DVI from parameter Sets A-C were analyzed together using a polynomial contrast to assess for dose-response. One subject completed one of the same IAPS forms twice inadvertently; the second administration is not included in the analysis.

## Results

### Safety

Subjects were prospectively queried at each session for any discomfort or pain due to the stimulation and were asked about any adverse events, as those reported in[21], since their last stimulation session. Across all 10 subjects and stimulation paradigms, there were no reports of any subjective distress or noticeable changes in cognition, emotional regulation, pain, or physical comfort[21].

In addition to a lack of subjective safety findings, subjects showed no objective safety risks either. They did not report any thermal changes at the stimulation site. No signs of edema, hemorrhage, or other structural disruption were observed on any images, and there were no changes in T1-weighted, T2-weighted, susceptibility-weighted, or diffusion-weighted images when compared to the previous session’s scan, as assessed by a board-certified neuroradiologist.

Across all study sessions, structural MRI demonstrated no significant interval changes. As no such changes were detected, only the initial and final scans required evaluation. Diffusion-weighted imaging was not available for review; however, ADC maps were assessed. In several cases, one ADC map per session showed an artifact that is typically associated with multiband acquisition.

Incidental findings included:

- Sub02: Punctate T1 hyperintense foci along the superior cortical surfaces, stable across sessions 1 and 5.
- Sub04: A small right frontal white matter T2 hyperintensity, nonspecific in appearance.
- Sub05: A ∼9 mm cyst in the right cingulate gyrus.

These incidental observations were stable, already present at baseline (i.e., prior to any sonication), and not considered significant by the reviewing neuroradiologist within the context of the study. No evidence of edema, hemorrhage, or other structural abnormalities was observed on T1-weighted, T2-weighted, susceptibility-weighted, or ADC map sequences.

### Modeling

Acoustic modeling indicates that the right amygdala was reached with tFUS amongst the participants (see Fig. 1 and Table 2). Distortion effects from the skull and other tissue types in the head shifted the focus of the ultrasound beam from the target region (usually towards the skull), this was only by a few millimeters in any direction.

**Table 2:** Estimated acoustic effects.

| Set | Sub | Sess | $I_{sppa.0}$<br>(W/cm<sup>2</sup>) | $I_{sppa}$ brain<br>max<br>(W/cm<sup>2</sup>) | $I_{sppa}$<br>target<br>(W/cm<sup>2</sup>) | $I_{spta.0}$<br>(W/cm<sup>2</sup>) | $I_{spta}$ brain<br>max<br>(W/cm<sup>2</sup>) | $I_{spta}$<br>target<br>(W/cm<sup>2</sup>) | MI | Temp |
| --- | --- | --- | --- | --- | --- | --- | --- | --- | --- | --- |
| A | 5 | 1 | 1.33 | 0.21 | 0.12 | 0.93 | 0.15 | 0.09 | 0.10 | 37.03 |
| B | 11 | 2 | 10.63 | 1.40 | 0.47 | 7.44 | 0.98 | 0.33 | 0.26 | 37.26 |
| C | 3 | 1 | 18.60 | 3.34 | 0.88 | 13.02 | 2.34 | 0.61 | 0.41 | 37.53 |
| D | 8 | 1 | 261.97 | 38.85 | 12.98 | 13.02 | 1.94 | 0.65 | 1.39 | 37.42 |

$I_{sppa.0}$ and $I_{spta.0}$ are the non-derated values output by the drive system. " $I_{sppa}$ target" and " $I_{spta}$ target" are the values at the target coordinate (a single voxel-space coordinate) defined in the individual T1-weighted image using the neuronavigation system for tFUS aiming. " $I_{sppa}$ max brain" and " $I_{spta}$ max brain" are the maximum values achieved within the focal point of the simulated ultrasound beam. "MI" and "temp" are the maximum MI and maximum temperature in the brain. Abbreviations: MI, mechanical index; req, required; Sess, session.
\*We report the $I_{sppa}$ and $I_{spta}$ achieved at the target voxel, which is the single coordinate in voxel that was defined and aimed at during tFUS administration.

### Behavioral Outcomes

The IAPS behavioral task was used to measure changes in emotion as a result of stimulation. The eight forms of the task that were created were randomized and counterbalanced across sessions. A form of the task was administered before and after each stimulation session.

### Change in Directional Valence Index

While there was a general dose-dependent decrease in mean DVI across stimulation paradigms, the Friedman test revealed no significant overall effect of the stimulation paradigm on the DVI (χ²(2) = 2.00, p = 0.368), with a small effect size indicated by Kendall’s W = 0.111. Post-hoc Conover tests with Holm correction showed no significant pairwise differences between any stimulus pairs (all p > 0.50). The changes in DVI are plotted in Figure 2a.

**Figure 2:**
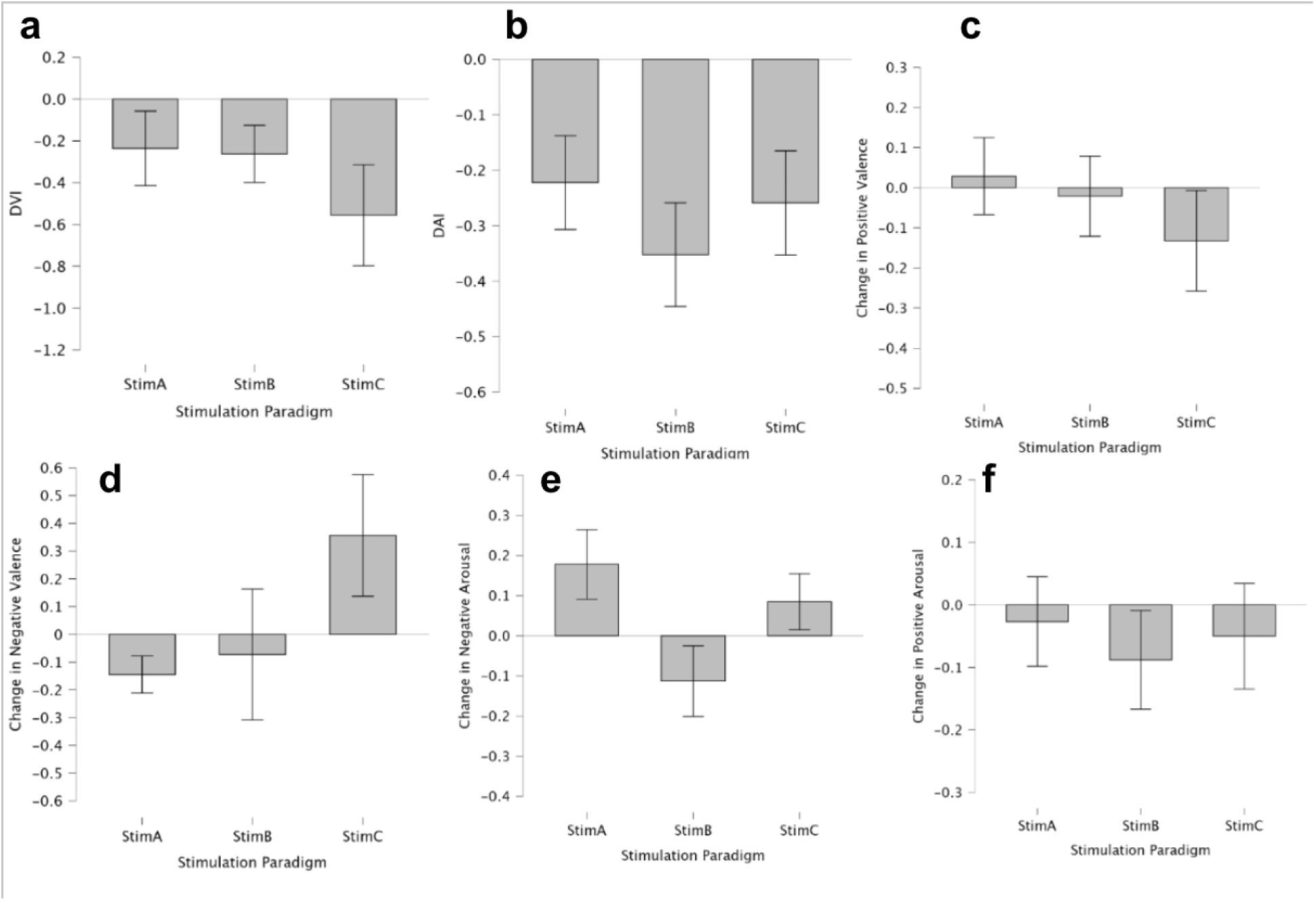
Behavioral Task Changes by Parameter Set. (a) Changes in DVI by stimulation paradigm. (b) Changes in DAI by stimulation paradigm. (c) Changes in positive valence by stimulation paradigm. (d) Changes in negative valence by stimulation paradigm. (e) Changes in negative arousal by stimulation paradigm. (f) Changes in positive arousal by stimulation paradigm.

### Change in Directional Arousal Index

The Friedman test revealed no significant overall effect of the stimulation paradigm on the DAI (χ²(2) = 0.941, p = 0.625), with a very small effect size indicated by Kendall’s W = 0.052. Post-hoc Conover tests with Holm correction showed no significant pairwise differences between any stimulus pairs (all p > 0.50). The changes in DAI are plotted in Figure 2b.

### Changes in Positive Valence

A repeated-measures ANOVA found no significant differences in positive valence changes across stimulation paradigms, F(2,16) = 0.585, p = 0.569, with a small effect size of η^2^ = 0.068. Post-hoc comparisons with Holm correction showed no significant differences between stimulus pairs (all p = 1.000), although the comparison between paradigm A and C revealed a moderate effect size (Cohen’s d = 0.53) despite being non-significant. Polynomial contrast did not show a significant linear (p= 0.352) or quadratic (p= 0.809) effect. The changes in positive valence by stimulation paradigm are plotted in Figure 2c.

### Changes in Negative Valence

A repeated-measures ANOVA found no significant differences in negative valence differences across stimulation paradigms, F(2,16) = 2.04, p = 0.163, η²_p_ = 0.203. However, polynomial contrast showed a significant linear trend was observed across paradigms A to C (p = 0.025), suggesting a possible dose-response pattern. The post-hoc Conover test with Holm correction between paradigm A and paradigm C approached significance (p = 0.075) and yielded a large effect size (d = 0.97). The changes in negative valence by stimulation paradigm are plotted in Figure 2d.

### Changes in Positive Arousal

The repeated-measures ANOVA did not show a significant effect of stimulation paradigm on the change in positive arousal (F(2,16)=0.16, p=0.858) with a small effect size of η_p_²= 0.019. Polynomial contrast did not show a significant linear (p=0.838) or quadratic (p=0.623) effect. Post-hoc pairwise comparisons with Holm correction showed no significant differences between any condition pairs (all p = 1.000). The changes in positive arousal by stimulation paradigm are plotted in Figure 2e.

### Changes in Negative Arousal

The repeated-measures ANOVA showed trend-level significance between changes in negative arousal across stimulation paradigms (F(2,16)=3.28, p=0.064) with a moderate to large effect size of n²_p_ = .29. Post-hoc comparisons using Holm correction revealed no significant pairwise differences between conditions (all p > .170). Polynomial contrast analysis indicated a trend-level significant quadratic trend across conditions (p = .053, d = 0.80), suggesting that changes in negative arousal may follow a quadratic pattern across stimulation paradigms. The changes in negative arousal by stimulation paradigm are plotted in Figure 2f.

### Directional Effect of Inhibitory versus Excitatory Stimulation

#### Directional Valence Index

The results revealed no significant difference in DVI between the two conditions, t((9) = –0.452, p = .662). These results suggest that, despite opposite presumed neuromodulatory directions (excitatory vs. inhibitory), there was no statistically detectable difference in directional valence modulation between paradigm C and D. The DVI plotted against the Stimulation Paradigm is seen in Figure 3a.

**Figure 3:**
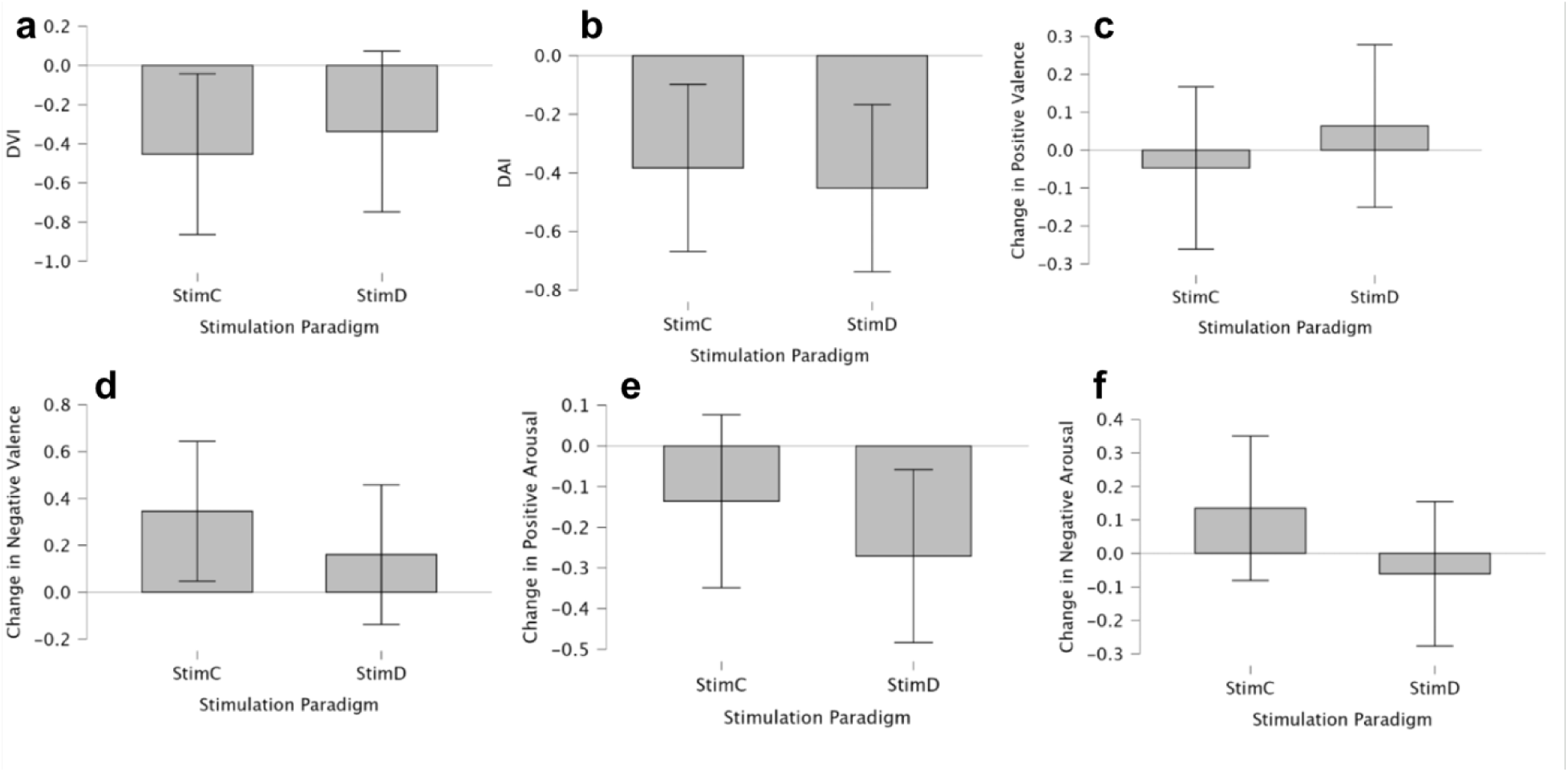
Directional Effect of Inhibitory versus Excitatory Stimulation on Behavior Task. (a) DVI by stimulation paradigm. (b) DAI by stimulation paradigm. (c) Changes in positive valence by stimulation paradigm. (d) Changes in negative valence by stimulation paradigm. (e) Changes in positive arousal by stimulation paradigm. (f) Changes in negative arousal by stimulation paradigm.

#### Directional Arousal Index

The DAI did not show significance on a paired t-test between Stimulation Paradigm C and Stimulation Paradigm D (t(9)= 0.387, p= 0.708). The DAI plotted against stimulation paradigm is seen in Figure 3b.

#### Change in Positive Valence

The change in positive valence did not show significance on a paired t-test between Stimulation Paradigm C and Stimulation Paradigm D (t(9)= -0.828, p=0.429). The change in positive valence plotted against Stimulation Paradigms C and D is seen in Figure 3c.

#### Change in Negative Valence

The change in negative valence did not show significance on a paired t-test between Stimulation Paradigm C and Stimulation Paradigm D (t(9)= 0.993, p= 0.347). The change in negative valence plotted against Stimulation Paradigms C and D is seen in Figure 3d.

#### Change in Positive Arousal

The change in positive arousal did not show significance on a paired t-test between Stimulation Paradigm C and Stimulation Paradigm D (t(9)= 1.106, p= 0.336). The change in positive arousal plotted against Stimulation Paradigms C and D is seen in Figure 3e.

#### Change in Negative Arousal

The change in negative arousal did not show significance on a paired t-test between Stimulation Paradigm C and Stimulation Paradigm D (t(9)= 1.454, p= 0.180). The change in negative arousal plotted against Stimulation Paradigms C and D is seen in Figure 3f.

## Discussion

This dose-escalation study demonstrated the initial safety and practicality of applying tFUS to the right amygdala in healthy volunteers, using intensities well above the FDA diagnostic output limit threshold for diagnostic ultrasound of I_SPTA.3_= 720 mW/cm^2^. Our primary safety aim was evaluated through structural MRI, specifically, whether any of the stimulation parameter sets would create changes on structural MRI. This means of safety evaluation was employed to mirror that used clinically during high-intensity focused ultrasound (HIFU) procedures, during which structural MRI is used to monitor for edema, hemorrhage, or unintended tissue damage during and after sonication[30]. Importantly, across all stimulation sessions, there were no structural changes observed on any MR scans. This held true even for the previously unused in-vivo stimulation conditions (Parameter Set C and D) with an I_SPTA.3_ of 10.08 W/cm^2^, as they did not lead to any adverse effects, despite the delivered intensity being 14 times higher than the FDA diagnostic limit. Additionally, MRI scans of patients who received Parameter Set D, which has a Mechanical Index of 2.8 (exceeding the iTRUSST recommended maximum of 1.9 [31]), did not show any of the structural changes, such as edema or microhemorrhage, that are typical with HIFU-like stimulation. Furthermore, there was a lack of any adverse events or serious adverse events during any stimulation session across subjects.

Together, these results show a level of safety within the parameter space used in our study. Although our sample size was small, with ten subjects, these findings still suggest the safety and practicality of these parameters for in vivo neuromodulation studies. They also raise questions about the current FDA intensity guidelines for I_SPTA_, set at 720 mW/cm². Given the the evidence from this study and others indicating safe tFUS intensities well above this limit, these findings suggest MI limits set by iTRUSST for therapeutic ultrasound may warrant reconsideration. Naturally, more robust future studies are needed before any official changes are made, but these results point to a potential need for reconsideration. Such adjustments might also help facilitate the approval process for future studies within this parameter space, possibly making IRB approval easier to obtain, and allowing for faster study initiation and smoother translation of innovative tFUS protocols to clinical applications. Additionally, understanding ultrasound targeting using techniques such as magnetic resonance acoustic radiation force impulse imaging (MR-ARFI) also requires higher stimulation intensities [32].

In addition to supporting the structural and regulatory safety of higher-intensity tFUS, our findings also offer preliminary insights into its temporal profile, raising important questions about the duration and persistence of neuromodulatory effects following stimulation. Even with sessions spaced one week apart, there was no cumulative or residual effect on emotional regulation, reactivity, or baseline emotional state, as measured by pre-stimulation behavioral task results, indicating that, even with significantly higher-intensity tFUS, neuromodulatory effects wore off between stimulation sessions. It is likely that with 10 subjects, we did not have sufficient power to detect an effect. While this further supports the temporal safety of this parameter space, it also points to an incomplete understanding of the temporal effects of tFUS neuromodulation. The precise duration of neuromodulatory effects induced by tFUS remains to be fully characterized, although prior studies in rodents and humans have demonstrated that such effects can persist for days to weeks[23, 33], underscoring the need for continued longitudinal evaluation within this new parameter space. Thus, while these results do demonstrate the safety of this parameter space and point towards further clinical trial testing, we are still unsure of the relationship, if any, between tFUS intensity and transient neuromodulatory effects.

The primary efficacy aim was to test whether increasing the intensity of ultrasound would produce a dose-dependent modulation of affective responses. Regarding our dose-response hypothesis, the results were rather mixed and inconclusive. Across parameter sets A to C, a significant linear trend emerged in changes in negative valence, indicating that increasing tFUS intensity led to a progressive shift in emotional reactivity to negative stimuli, suggestive of a dose-response pattern. This pattern was further expected given the role of the right amygdala, which is traditionally associated with negative emotional processing[34, 35]. In parallel, across parameter sets A to C, there was a general dose-response decrease in DVI and positive valence, although these trends were non-significant. However, given the small sample size of this study, these changes cannot be definitively used as evidence of an intensity-dependent effect of right amygdalar tFUS on valence.

With respect to changes in arousal across paradigms A to C, the DAI and positive arousal changes did not rise to the level of statistical significance. Notably, there was a marginally significant quadratic trend identified across changes in negative arousal, indicating that the relationship between stimulation intensity and negative arousal may follow a non-linear pattern. This raises the possibility that intermediate stimulation levels may suppress negative arousal more strongly than lower or higher levels. However, while such non-linear dose–response curves have been described in other neuromodulation contexts, particularly in studies targeting subcortical structures like the amygdala[12, 36], it is more likely that the observed trend was a result of small sample size variability and methodological limitations of the behavioral task used in the study.

When comparing Stimulation Paradigm C, believed to be excitatory (10.08 W/cm^2^ I_SPTA.3_, 70% DC) and Stimulation Paradigm D, which is believed to be inhibitory (10.08 W/cm^2^ I_SPTA.3_, 5% DC), no metric reached a level of significance. It’s likely then that while these paradigms differ theoretically in their predicted neuromodulatory direction, the task used here was not sensitive enough to detect these specific effects, especially given the small sample size in this pilot study. Additionally, it’s possible that the modulation occurred at a neurophysiological level that did not lead to immediate directional changes in arousal or valence. As such, future work should ensure that concurrent neuroimaging or electrophysiology is collected to assess the polarity of the neuromodulation on a more granular level.

Collectively, these results provide some evidence for a dose-response effect of tFUS intensity, however, they are largely inconsistent and inconclusive. Beyond the limitations imposed by the small sample size, it is also important to note that this trial was conducted in healthy subjects, and as such, the primary endpoint was safety rather than therapeutic efficacy. The tasks used are primarily meant for anxious individuals, not for healthy volunteers. Subsequently, the lack of clear and robust neuromodulatory effects was not unexpected. We anticipate that installing these higher intensity tFUS parameters in clinical populations with dysregulated affective circuitry, such as in mood or anxiety disorders, could result in clearer effects, especially in the context of targeting abnormally hyperactive or hypoactive circuits. This is especially relevant given that prior studies were constrained by the FDA’s diagnostic ultrasound intensity limit of I_SPTA.3_ = 720 mW/cm² may have underdelivered energy relative to what is needed for clinically meaningful engagement. Ultimately, replication of these findings in patient populations, with symptom-specific targets and outcomes, will be essential to determine the true translational value of higher-intensity tFUS.

## Conclusion

This dose-escalation study provides preliminary evidence that low-intensity tFUS can be safely performed at intensities above the current FDA limit and was applied safely to the right amygdala in healthy volunteers. Across all stimulation sessions, structural MRI and behavioral monitoring revealed no adverse events, edema, hemorrhage, or other tissue disruption, even at intensities exceeding the current FDA diagnostic ultrasound output limit and the iTRUSST recommended maximum. These findings establish an empirical foundation for reconsidering whether such conservative regulatory thresholds are appropriate for neuromodulation research. While exploratory analyses suggested possible dose-dependent effects on affective processing, particularly in negative valence and arousal, these results were inconsistent and likely limited by small sample size and the use of healthy controls. The complete absence of adverse effects, together with preliminary signals of neuromodulatory engagement, supports the continued evaluation of higher-intensity parameters in clinical populations. Future studies with larger samples, patient cohorts, and concurrent functional neuroimaging will be essential for clarifying both the therapeutic potential and mechanistic specificity of tFUS beyond current FDA and iTRUSST limits.

## Data Availability

All data produced in the present study are available upon reasonable request to the authors.

